# A Qualitative Expert Interview Study: Mobility Health in Indonesia

**DOI:** 10.1101/2021.12.13.21267758

**Authors:** Mikhael Yosia, Ray Wagiu Basrowi, Tonny Sundjaya, Bertri Maulidya Masita

## Abstract

**Background:** Indonesia has an ageing population that can develop mobility health-related problems in the future, including osteoporosis, arthritis, sarcopenia, low back pain, neck pain, and peripheral neuropathy. However, there are limited references and research that look upon mobility health and problems surrounding it in Indonesia.

**Aims:** To improve understanding on issues surrounding mobility health in Indonesia through a semi-structured interview with relevant experts.

**Method:** Semi-structured qualitative interviews via phone calls were conducted with eight different experts. Data were analysed using reflexive thematic analysis. Experts with experience dealing with mobility (bone, muscle, joint, movement) related issues for more than ten years, working in Indonesia, and communicating in English or Bahasa Indonesia were included.

**Results:** Four themes were then identified across the experts on issues surrounding mobility health in Indonesia; this includes [1] screening and assessment of mobility health, [2] treatment for mobility health problems, [3] awareness campaign, [4] supplement for mobility health.

**Conclusions:** Diagnostic modalities for mobility health are abundant but still expensive. Treatment of mobility health with herbal medicine (*jamu*) might be beneficial. Fortified food or milk can improve mobility health. Social media can be a promising tool to increase awareness regarding mobility health. In general, there needs to be a change in mindset from curative to prevention in both health care providers and the general population.

## INTRODUCTION

Passage of time undoubtedly caused gradual structural and functional changes in any organism. The process of ageing is inevitable and often causes deteriorating physical functions until the time of death. Ageing is also evident in the decrease of the body’s ability to perform physical movement following the deterioration of musculoskeletal and neuromotor functions due to a decrease in joint’s flexibility, loss of muscle mass and reduction in bone mineral density^1,2^.

If left unchecked, combined with the right conditions, the deterioration resulted in common mobility-related diseases; arthritis, osteoporosis, sarcopenia, low back pain, neck pain, and peripheral neuropathy^3,4^. The decline in mobility undoubtedly caused a significant decrease in an individual’s independence and overall quality of life, represented by the finding by the Netherland’s *Centraal Bureau voor de Statistiek* (CBS) that 30% of elderly with mobility problems complain that they are less content with life and 25% generally feels less happy with their current condition^5^. With the prevalence of mobility-related problems in the elderly reaching up to 35% at 75 years of age^6^, this issue should not be underestimated.

The term “mobility health” is uncommon in daily clinical practice, yet it is interchangeable or mostly covered in musculoskeletal and neuromotor well-being services. Although the awareness of mobility health is lacking, the problem will only accentuate in the future. As the fifth most populous country, Indonesia is estimated to have 30% of its population at 50 and above, which translates to around 82 million people with a potential mobility issue^7^. However, finding basic information such as the prevalence of musculoskeletal problems in Indonesia is lacking at best. With the absence of evidence and references on mobility health in Indonesia from qualitative and quantitative studies, any revelation on this issue will be valuable for healthcare practitioners and the general public.

This paper aims to improve understanding on issues surrounding mobility health in Indonesia through a semi-structured interview with relevant experts. The interview will be conducted to understand key issues surrounding mobility in Indonesia, identify triggers to improve awareness of mobility health in adults and the elderly and highlight opportunities to improve mobility health in Indonesia. With unique cultural and religious aspects playing a significant role in Indonesian’s health-seeking behaviour^8^, a qualitative study is hoped to reveal the nuance of mobility health and how it is perceived in Indonesia.

## METHODS

### Design

This study is qualitative empirical research through an expert interview that was conducted in July 2020. The interviews were used to gather expert’s knowledge on mobility health whereby the “participant is attributed as an expert by virtue of his role as an informant”^9^. The experts are considered people with experience in a certain field or possess information, training, or education not accessible to others. The study involves a semi-structured phone interview with eight mobility experts from different fields in Indonesia. Semi-structured interview is chosen as it may shed valuable knowledge from participants while keeping on the topic of interest^10^. Prior to the interview, the authors made guiding questions and then discussed, and pilot tested the list of questions to a research member with a background in mobility health.

A list of medical experts, influencers, religious leaders, and fitness experts on mobility is gathered before shortlisted based on the convenience sampling technique. Participants who are available, willing, could converse in Indonesian/English and had access to telephones are included in the in-depth interview. Each interview session lasted for about 30-40 minutes, and the interviewer was guided with a set of questions to keep the conversation relevant to the study’s purpose. If required, participants were encouraged to add any additional relevant information.

Writing of this report were then made based on the Consolidated Criteria for Reporting Qualitative Research Checklist (COREQ) guidelines^11^. The study was approved by the Ethical Review Board of Faculty of Medicine, Universitas Atma Jaya (NO: 13/11/KEP-FKIKUAJ/2020).

### Methodological Approach and Participants

Through in-depth semi-structured interviews of relevant experts in mobility, this paper aims to shed more information on mobility and its related problem in Indonesia. This includes gathering insight on the current status, patient pathway, patient behaviour, and trends on mobility health in Indonesia through experts who worked directly with patients with mobility problems in their daily practices. The interviews will also be utilised to understand the expert’s perceptions on key priorities surrounding mobility health and whether there is a gap in achieving these priorities. Further information will be gathered to identify other challenges and recommendations to improve Indonesia’s early awareness, compliance, and nutritional support for mobility. Other information gathered from the literature search will also be cross analysed with opinions from these experts.

A list of experts was gathered through internet research (Google and social media), word of mouth, and other relevant mobility experts were identified based on references from other experts. For this study, experts’ eligibility was based on the following inclusion criteria: had been working or had experience dealing with mobility (bone, muscle, joint, movement) related issues for more than 10 years, working in Indonesia, and communicating in English or Bahasa Indonesia. Self-proclaimed experts dealing with what is thought to be pseudo-medical science (spiritual healer, a practitioner of unorthodox treatments, etc.) were excluded as part of the study as information gathered is thought to be biased by their non-evidence-based practice. None of the experts approached refused to be part of the study. The eight final experts were all health care providers in Indonesia with expertise in mobility; this includes experts with profiles as social media influencers, religious leaders, and fitness experts. A full list of the participants can be seen in Table 1.

**Table 1.**
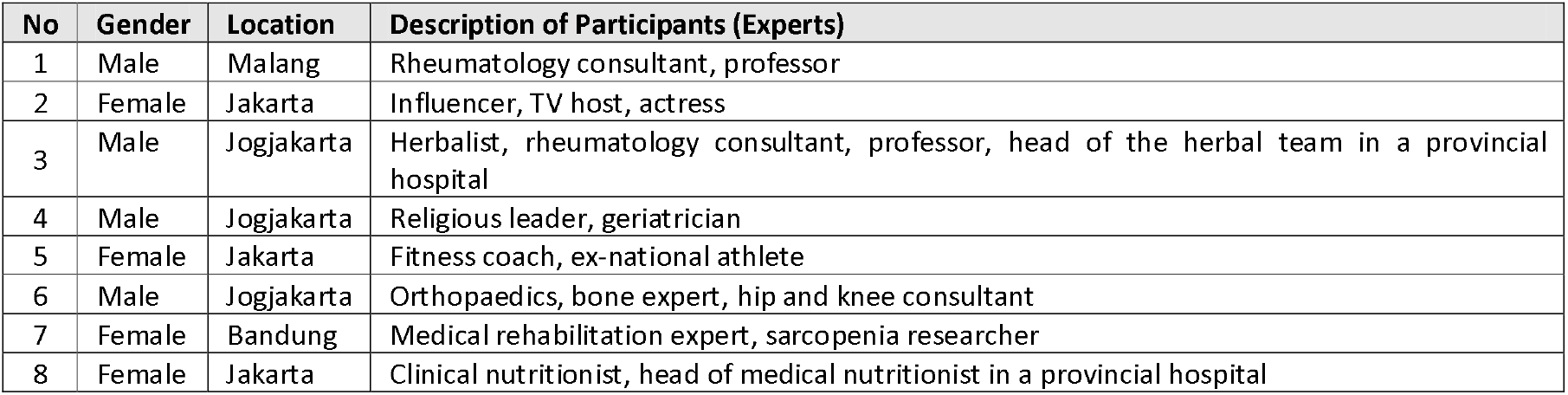
The descriptions of participants invited to be interviewed as experts for this study. Do note that all the participants are health care providers.

After considering the pros and cons, the interviews were conducted via phone calls, which was chosen because of flexibility in timing, geography (to avoid clustering all the experts in one geographical location, e.g. in the capital, Jakarta), and for the sake of practicality amidst COVID-19 pandemic^12^. The researchers are fully aware that using asynchronous communication may reduce the perception of social cues or non-verbal responses towards the questions. Using a non-face-to-face method would also mean that there are challenges in creating a conducive interview ambience; an expert might be doing other work during the interview or being called away to see a patient, all of which may disrupt the interview flow^13^. Although it is impossible to eliminate the issues, the researcher took steps to minimize these disruptions, including arranging the call based on the availability of the experts and agreeing upon the best time to conduct the discussion. This arrangement would also ensure that the experts had ample time to reflect on their thoughts, opinions and feelings for the interview.

After the interviews, an internal discussion between the researchers was made to determine whether additional experts needed to be interviewed. Since a vast range of insight and information had already been gathered, the researchers agree that the number of experts is sufficient to achieve the aim of this study. No repeat interviews were conducted with any of the experts. All the information gathered were then analysed and written into the final manuscript.

### Procedure

Due to the COVID-19 pandemic, all expert’s interview was done via phone calls. Electronic mail was sent to promising candidates with information on the study and method of the interview should they be willing to participate. The experts were instructed to reply to the message if they consented, followed by arranging a phone call appointment. The objective of the individual interviews is to collect insights from individual experts on mobility health that will be analysed and consolidated by research teams. The individual interview was set to minimise the external influence on interviewees’ answers, common in group interviews.

Prior to and after the interview, no materials about mobility health were provided to the experts. The target of each interview was to conduct a true natural discussion where the interview speaks freely but is oriented by open-ended guiding questions. Research members with a background in medical health conducted all the interviews.

The interviews were separated into three main parts:

1. Introduction – Introduction from the interviewers, explanation of the study, and their involvement are completely voluntary, followed by the confidentiality of the discussions.
2. Questions - A set of predetermined open-ended questions were given to guide the interview to keep the discussion relevant.
3. Conclusion – Experts are encouraged to share their thoughts on anything related to mobility health, followed by a recap and summary from the discussion.

Key insights from the interview were written down verbatim and used as quotes to represent the expert’s opinion on a certain issue; these were then repeated to the experts to ensure that it had captured the point. The interviewer wrote down a summary, and key points gathered during and right after the interview. All the interview notes and quotes were kept in a Word document.

### Reflexivity

The interview was conducted by a male researcher with vast experience as a research consultant specialising in innovation in the healthcare and food industries. The interviewer is a PhD, MBA, and a Bachelor of Science from reputable universities who had a great understanding of conducting qualitative research prior to this study. The interviewer is familiar with Good Clinical Practice (GCP) and ethical research following the Declaration of Helsinki.

Prior to the interview, no relationship was established between the interviewer and the interviewee. The participants had not been given any personal information or motivation on why the interviewer was conducting the research. The interviewer was not aware of any personal information apart from the description of participants available in Table 1. Throughout the interview, the interviewer tried to be aware of personal beliefs and assumptions that the expert had on mobility health and to avoid including these assumptions in the analysis.

### Analysis

Expert’s identifiers (full name, titles, contact number, work address) were not included during the interview and note-taking process (in Microsoft Words). Experts are addressed based on the number and descriptions of their work, as seen in Table 1. A reflexive thematic analysis approach was used on the data. The researchers gathered and went through the notes to search for patterns and meanings in the conversations. Quotes and notes were then labelled and organized into groups, creating themes. The researchers then review the notes to ensure enough data is present to support each of the themes. After data and main themes had been identified, the researcher organized it into an argument that suits the aim of this study.

## FINDINGS

Throughout the expert interviews, key pieces of information were gathered around the issue of mobility in Indonesia. The expert main concerns and focus expressed during the talk are summarized in Table 2. Below are excerpts from the interviews with all the experts separated into several key topics.

**Table 2.** Experts interest and focus in healthcare for mobility health.

| Expert | Level of Interest |  |  |  | Highlight |
| --- | --- | --- | --- | --- | --- |
|  | Prevention | Screening | Treatment | Compliance |  |
| 1 | Medium | Low | High | Med | Glucosamine or chondroitin are not recommended for joint's health due to lack of evidence; lifestyle change for joint treatment is key. |
| 2 | High | Medium | Medium | High | People focus on curative, not preventive. TV is the top media in populations above 50 years old for awareness and compliance of supplements. |
| 3 | High | Low | High | Medium | Herbal medicine or jamu is used as a complementary treatment and may reduce the dose of modern treatment. |
| 4 | High | High | High | Medium | Religious activity (prayer/gathering/pilgrim) can be a source of awareness, screening and consideration on treatment for mobility. |
| 5 | High | High | Low | High | Screening should be related to daily activity to increase relatability, as this is important for compliance. |
| 6 | High | Medium | High | Low | The habit of drinking milk should start from a young age (<35 years old) to build enough bone mass to prevent osteoporosis. |
| 7 | Medium | High | High | High | Mobility screening tests can increase awareness of mobility problems. Sports community and friends are key for improving compliance. |
| 8 | High | Low | Low | Medium | People needs to be aware of nutrition recall to increase awareness of their nutrition intake and take necessary preventive steps. |

### 1. Screening and Assessment of Mobility Health

#### 1.1. On general assessment of mobility health

General evaluation of mobility includes evaluation of strength, balance, endurance, and functionality, but these tests cannot pinpoint the aetiology of any mobility-related issues. The experts mentioned several general mobility assessments or screening tests available and commonly used in Indonesia (see Table 3).

**Table 3.**
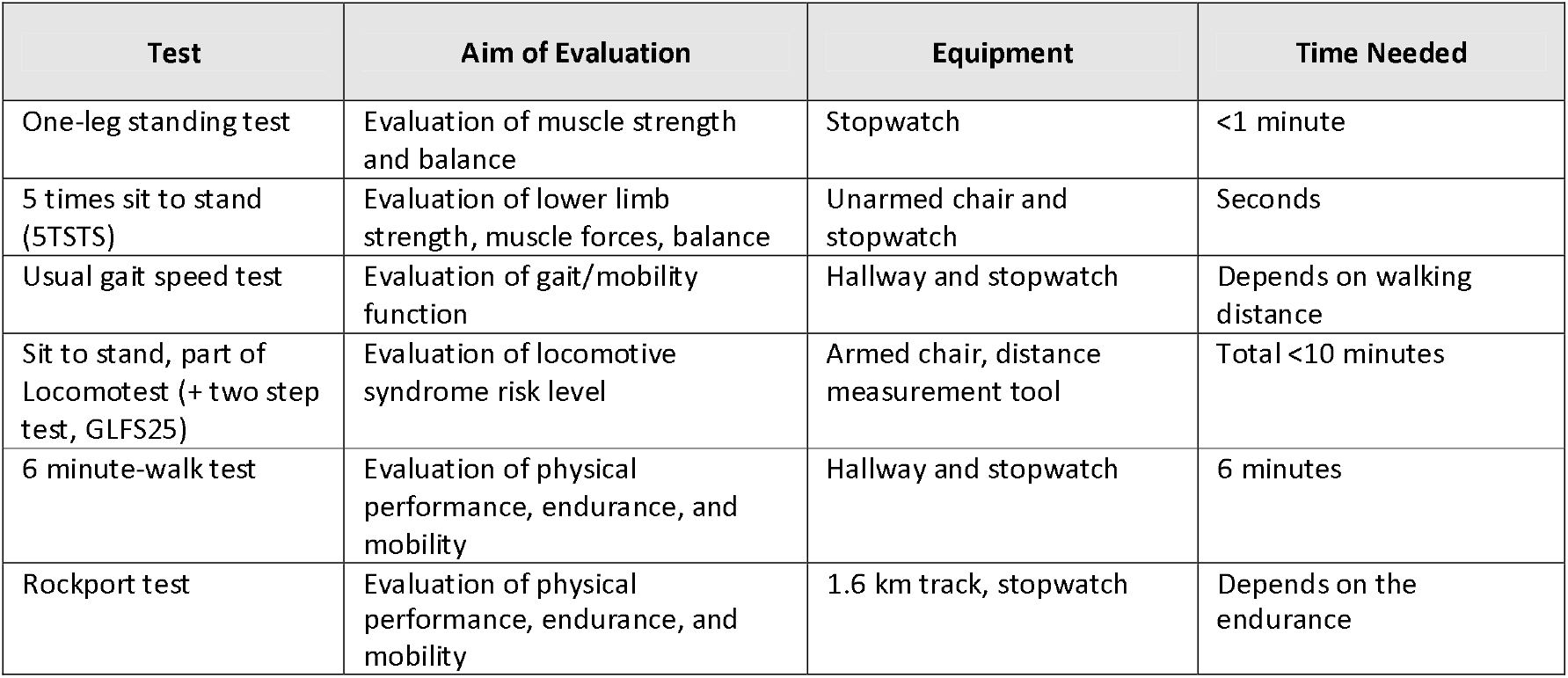
General mobility assessment or screening test mentioned by the experts.

*“These are valid functional test on mobility; the inability to have certain value or reaching a value below the average indicate that there is functional issues on mobility, however, these tests cannot pin-point the specific causes of the problems, for example: which specific parts and function of the body is affected? Ankle, knee, muscle, strength, balance, flexibility*.*”*

*(Expert 7, Medical rehabilitation expert)*

The experts had also mentioned that other than mobility, some of these tests are good at determining an individual’s level of fitness or endurance. These general assessment tests are good at raising awareness of mobility health among the general population. However, it is not recommended for clinical screening or diagnostic as these requires a further physical examination, specific test, and regular check-up and follow-up.

#### 1.2. On screening for specific organ or mobility issue

In order to do a clinical assessment of mobility issues, there are a series of specific assessments, which includes a physical examination, radiography, or lab test. The experts had identified several specific tests for common mobility issues in Indonesia; arthritis, sarcopenia, osteoporosis (see Table 4).

**Table 4.** Specific diagnostic test available for clinical screening and diagnosis of arthritis, sarcopenia, or osteoporosis.

|  | Joint - Arthritis | Muscle - Sarcopenia | Bone - Osteoporosis |
| --- | --- | --- | --- |
| **Without equipment** | - Risk factor questionnaire<br>- No reliable tools for early screening, need holistic check on joint | - Risk factor or protective factor questionnaires<br>- Screening with SARC-F: lifting weight, assistance in walking, rise from chair, falls | - Risk factor or protective factor questionnaires – SCORE, FRAX, OST |
| **Simple test** | - Uric acid strip test for gout arthritis (similar to glucose/cholesterol test) | - Usual gait speed<br>- Hand gripping strength<br>- Upper arm and calf circumference over time | - None readily available in Indonesia |
| **Complex test** | - Joint radiography<br>- Lab test for markers of joint diseases | - Bioelectric impedance in gym/hospital<br>- DXA (Dual energy X ray absorptiometry) only available in around 34 hospitals in Indonesia | - Ultrasound<br>- DXA test in hospital (Gold standard, around IDR 700,000)<br>- Lab test for bone mass density (around IDR 700,000) |

*“It is expensive to check for vitamin D level in blood or bone mass density (around IDR 700,000 per test), which may reduce the awareness of vitamin D deficiency and osteoporosis*.*”*

*(Expert 8, Clinical nutritionist)*

Although there are options available for specific testing of mobility-related diseases, experts think that the test’s specificity and sensitivity can be increased by determining a reference or cut of value specific for the Indonesian population. The price of several complex tests is quite expensive, which deters people from screening (or even getting a diagnosis) for mobility diseases. It should also be noted that some of the tests (SARC-F) have qualitative scoring, which relies on an individual’s subjective judgement, which may skew the result.

#### 1.3. Provision of screening for awareness campaign

Provision of free screening (e.g. BMD measurement using ultrasound or body composition screening using bioelectric impedance analysis) are sometimes offered in public places or events to increase awareness on mobility health.

*“In terms of the emotional aspect, screening may incite awareness and sometimes fear when the screening results shows a bad result, this somewhat forced people to do exercise or change to a healthy diet, but they may not be able to enjoy the changes through subsequent test result which make this one time free screening unsustainable in the long term*.*”*

*(Expert 5, Fitness coach)*

#### 1.4. Utilizing digital tools for screening or awareness campaign

Nowadays, the utilization of apps and IT for medical purposes has significantly increased. Digital tools for early screening include questionnaires with questions on an individual’s history and physical assessment, while prevention and monitoring tools include mobile applications for health challenges, health tracking and education.

Although promising experts think that digital tools may not bode well with the elderly (population above 50 years of age) because of unfamiliarity in the majority of the elderly population with utilizing apps in mobile phones, while the health challenges for elderly with limited mobility may not be interesting and even demotivates them as they struggle to complete the challenges. It may be better to relate the challenges and screening points to relatable daily activities (e.g. lifting luggage while travelling, inability to move during a social gathering, or conducting prayers) to incite emotional connections.

### 2. Treatment for Mobility Health Problems

#### 2.1. Herbal medicine for mobility health

In Indonesia, traditional treatment and herbal medicine (*jamu*) are sometimes used alongside standard clinical treatment. The experts state that most health care providers allow the patient to have *jamu*, massages, and acupuncture treatment when it meets their guidelines and standard (e.g. traditional treatment corresponds with the diagnosis).

*Jamu* is commonly used as a complementary treatment (see Table 5 for a list of *jamu* and its uses) and, in some cases, were used to reduce the dosage of modern medicine. There is a widespread belief that traditional herbal medicine is “better” than modern medicine, with the expert stating that nearly half of the adult population prefer traditional herbal medicine to help ease mobility-related health problems. There is also a preference from an expert that patients prepare their *jamu* at home instead of purchasing through an unlicensed store/home industry as they usually add steroids to increase the *jamu*’s “potency”.

**Table 5.** List of *jamu* and herbal ingredients and it’s effect to mobility health.

| Joint | Muscle | Bone |
| --- | --- | --- |
| - Anti-inflammatory, antioxidant: ginger, curcuma, turmeric, sambiloto, brotowali, and Black cumin<br>- Anti-hyperurisemia for gout arthritis: sidaguri, celery, bay leave<br>- The general public believe in kolang kaling (sugar palm fruit) but there is no evidence yet | - Jamu /herbal are common for muscle pain, due to it’s anti-inflammatory, analgesic, and antioxidant properties: beras kencur (beras sangria, ginger, aromatic ginger, turmeric), cabe puyang (cabai jawa, lempuyang, aromatic ginger)<br>- Evidence based herbal used to increase muscle strength and muscle mass is pegagan (Indian pennywort) | Anti-osteoporosis herbal medicine that has been proven to influence the bone formation or resorption metabolism: cloves, cinnamon, sambiloto, great morinda, onion, mung bean. |

Traditional medicine such as *jamu* (herbal medicine), acupuncture, and massages had been incorporated in the public health services, with provincial hospitals creating specific outpatient clinics for these services. Expert 3 (Herbalist) states that even though these services are available, there are few patients because traditional medicine is not covered by the national public insurance (BPJS). The doctor must first diagnose patients who ask for herbal treatment before a matching treatment is administered.

*“Choices of Jamu for mobility issue is based on popularity and influenced by the taste, some people do not like Sambiloto, Brotowali because they are very bitter*.*”*

*(Expert 3, Herbalist)*

#### 2.2. Compliance on treatment and management

Compliance with assigned treatment is a multifaceted issue with no silver bullet. According to the experts, difficulty in compliance is caused by the patient’s behaviour and lifestyle and economic reasons related to the price of treatments. Patient with higher socio-economic status usually has better awareness and knowledge, thus a higher compliance rate. Sustaining lifestyle changes is difficult because patients tend to stop their diet, assigned physical exercise, and medication when their complaints subside, or simply because they are “bored”. Some patients still fear continuous use of medicine because they think the medications are “toxic” if consumed for an extended period. The expensive price of some medication and treatment also caused compliance problems, with patients refusing to continue with their assigned treatment if it is not covered by the national health insurance (BPJS).

*“The compliance on treatment or lifestyle change is always an issue, and one of the ways to make them comply is through involvement and reminder from their family or caregiver*.*”*

*(Expert 1, Rheumatology Consultant)*

The expert thinks that the provision of campaigns on television and social media regarding nutrition and healthy lifestyles may improve compliance with lifestyle changes. Involvement of patient’s friends/social groups (e.g. religious, neighbourhood community) in regular exercise activity may improve the patient’s willingness to continue with their treatment. It is also important to pay more attention to the caregivers, as they remind the patients of their assigned treatment and management, which naturally would increase compliance.

#### 2.3. Gap in patient care for mobility health

The expert had raised gaps in the prevention and treatment of mobility health problem that may be beneficial if addressed properly (see table 6).

**Table 6.** Summary of gaps present in care for mobility health problems

| Joint | Muscle | Bone |
| --- | --- | --- |
| **Gap in early screening for joint health**<br>- There is no reliable early screening to improve awareness of joint health<br>- Early diagnosis is only available through radiography and laboratory test, which is | **Lack in early awareness of muscle loss with ageing**<br>- Most people are not informed about sarcopenia or muscle loss<br>**There is no established standard drug available for treatment of muscle loss**<br>- Most drug treatments available in | **Even though there's sufficient awareness in bone problems, there are no concrete action for long term prevention.**<br>- The mindset of general populations are still on curative rather than preventive<br>**There is no simple early screening** |
| expensive<br>**Gap for a strong evidence of supplement for joint health**<br>- Glucosamine and chondroitin are not strongly supported by scientific evidence in improving the metabolism of bone and cartilage<br>- Some professionals do not recommend glucosamine and chondroitin for joint supplements | Indonesia are still under investigation | **method for osteoporosis**<br>- The early screening needs equipment for bone mineral density measurement, mostly performed in healthcare facility |

*“There is no strong evidence that a specific drug is effective for muscle loss treatment, we usually use exercise and nutrition to improve the muscle mass/strength”*

*(Expert 7, Medical rehabilitation expert)*

*“There is no simple early screening method for osteoporosis because it is silent disease; DXA as the gold standard are only available in hospitals”*

*(Expert 6, Orthopaedics)*

### 3. Awareness campaign

#### 3.1. Media for awareness campaign

Public awareness for mobility health mainly comes from online or social media (messaging services, fitness/health app and platform, podcast), mass media (TV advertisement, talk shows), word of mouth (religious or social gathering, workplace, sports community), and offline events (seminar, conferences, etc.).

According to the expert, each age group had a different preference for media. Television still plays an important role in influencing people >50 years old in choosing supplements for mobility health. At the same time, social media (especially messaging groups) can spread viral information and disseminate facts or hoaxes on mobility health rapidly to those >50 years old. In the younger age group (35-45 years old), social media still influenced the spread of information on mobility health. Interestingly, friends and caregivers seem to have high influence via word of mouth in treatment choices and supplements for mobility health.

#### 3.2. Religious activity and mobility awareness

In Indonesia, religious activity and gathering can be a promising place to increase awareness for mobility health. As Indonesia had the largest Muslim population globally, the Muslim prayer movement, religious gathering and religious pilgrimage had surprisingly become a source of realization and awareness for mobility health. Most mobility health issues are recognized through discomfort during the Islam prayer movement, which involves kneeling, sitting, prostrating, and standing straight. There is also a health check conducted 1 year before the pilgrimage to Mecca; this includes screening to determine the would-be pilgrim’s fitness and mobility status (Rockport test, 6 min walk test, etc.), followed by a recommendation for regular weekly exercise to improve fitness and mobility health.

With the recognition of problems during prayers, religious events also became a hotspot for discussions and sharing of experience and knowledge on mobility health issues and treatment for those issues. Active religious congregants tend to use herbal remedies, which includes Islamic herbal treatment or jamu; it is common to see the preachers and congregants selling Islamic natural herbal treatment (honey, black cumin, dates, olive oil)

*“Preachers or religious leader who have background in the health sectors e*.*g. doctors, pharmacist, herbalist often include health and nutrition as topics in their preaching, but for non-health expert, there is not much involvement of nutrition or mobility health on their preaching*.*”*

*(Expert 4, Religious leader)*

### 4. Supplement for Mobility Health

#### 4.1. Public consideration on supplement and herbal supplement

According to the expert, standard supplements are popular for osteoporosis and osteoarthritis, while herbal supplements or a combination of both are popular to support treatment of muscle and joint (gout arthritis) pain.

*“Adult choose a supplement based on the popularity as compared to scientific evidence. Products with more campaigns on television/social media are typically chosen. Price also has a huge effect on choices*.*”*

*(Expert 2, Influencer)*

#### 4.2. Public consideration on fortified food product

Fortified food products in Indonesia focus on bone health, with less emphasis on muscle and joint health. The emphasis includes more promotion of bone health awareness (especially osteoporosis) in the advertisements. The general public tends to use standard or herbal supplements for joint and muscle problems.

*“Fortified milk for adult is mostly campaigned for bone and osteoporosis prevention, while there is less exposure for muscle loss/strength*.*”*

*(Expert 7, Medical rehabilitation expert)*

#### 4.3. Supplement purchasing behaviour

Indonesians concentrate more on curative rather than preventive, with the purchase of supplements mostly done to support the main treatment. See Figure 1 for a breakdown of supplement purchasing behaviour according to the experts.

**Figure 1.**
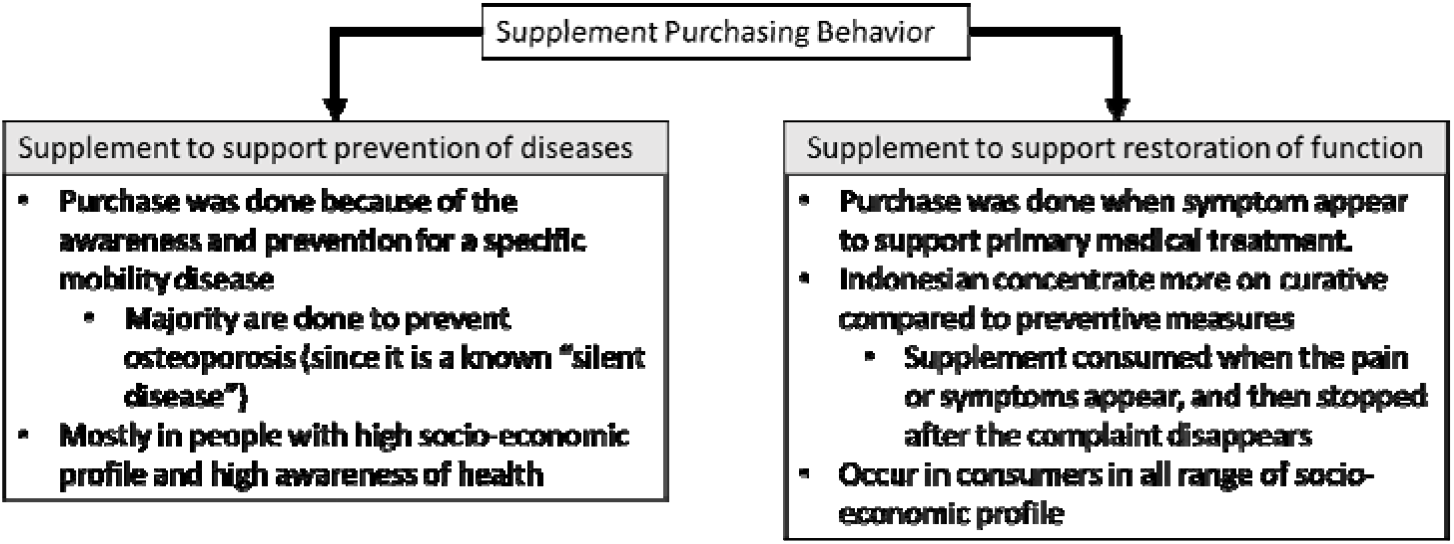
Summary of supplement purchasing behaviour in Indonesia

*“Doctors in public hospital mostly prescribe generic supplement e*.*g. Calcium, vitamin D for patient it is covered by the national insurance scheme (BPJS)”*

*(Expert 6, Orthopaedics)*

## DISCUSSION

This paper aimed to improve understanding of issues surrounding mobility health in Indonesia through a semi-structured interview with relevant experts. With an impending increase in the impact of mobility health and the lack of available evidence, this qualitative study is a good start for revealing the current status of mobility health in Indonesia. Through the semi-structured interview conducted with the eight experts, the researcher had determined four main topics of interest; [1] Screening and assessment of mobility health, [2] Treatment for mobility health, [3] Awareness campaign, and [4] Supplement for mobility health. Experts from different professional backgrounds related to mobility health were interviewed, which shed new information presented in this paper and helped increase understanding of the current status of mobility health in Indonesia.

### Screening and Assessment of Mobility Health

As most mobility health disorders are barely noticeable in the early stages, screening may help detect these issues earlier and increase the chance of patients receiving earlier treatment and a better prognosis^14^. General assessment on mobility is present in Indonesia and applied to the general public to a certain extent. Although available to indicate a person’s superficial mobility status, these general assessment does not give specific information on which target organ or function are affected. Simple test such as the one-leg-standing test, sitting-rising test, or sit-to-stand test may give general evaluation on muscle strength and balance, but should not be used alone as basis of clinical diagnosis^15,16^.

Even though most people will only seek screening tests after they had sufficient awareness of a certain disease^17,18^, due to its simplicity, these general tests can be conducted *en masse* at public gatherings involving many people, thus making it useful as an interactive means to increase awareness of mobility health. This potential is shown by the Locomo (sit-to-stand) challenge conducted in 2018 by Hakuhodo with the Japan Health, Labour and Welfare Ministry resulted in 1 billion media impressions and an unprecedented 80% increase in awareness towards locomotive syndrome in Japan^19,20^. With the COVID-19 pandemic in mind followed by increase of self-isolation and sedentary lifestyle; the utilization of these general tests has increased as it can be done independently at home guided by simple online instructions^21^, potentially reaching rural areas and increase awareness of mobility health around Indonesia.

Numerous advanced specific diagnostic examinations for mobility health is available in Indonesia to properly diagnose mobility health problems, but advanced technology comes at a high price. At times, a simple blood test (for vitamin D) or bone mass density test can still be considered expensive for most Indonesians. Even though all Indonesian are covered by the National Health Insurance (BPJS), its referral system and lack of advocacy on how to properly utilize the BPJS made it unattractive for certain people – those who are well off and able to afford private treatment or those with low socioeconomic status without enough knowledge and information on how to utilize the BPJS properly^22^. The addition of low awareness towards mobility health may accumulates to low utilization of these advanced diagnostic modalities.

The provision of free screening by a private company or the government is available in public places. However, as one of the experts’ notes, these events sometimes lack sustainability. Without the proper knowledge or resources to access a certain screening test, there is no continuity towards the monitoring of an individual’s mobility health status, rendering the one-off screening event rather impractical as it might not show the results of lifestyle changes that an individual had done. Interestingly, the experts also note that utilization of apps and IT in Indonesia for free screening is deemed ineffective for the elderly as most are not familiar with how to utilize the apps properly. A study that looked upon this issue had pinpointed that apps are usually designed to cater to the young; a user-centred design with a user-friendly interface for the elderly might help solve this issue, even though it would not remove the hesitancy of using smartphones or tablets altogether^23^.

### Treatment for Mobility Health Problems

With most Indonesian still consuming traditional medicine, *jamu* often becomes a treatment choice for mobility health issues. *Jamu* is traditional medicine usually made from spices and plants, and it has been known to have analgesic, anti-inflammatory, and antioxidant properties^24^. Throughout time, jamu has been extremely popular as an analgesic used in reducing complaints of joints, muscle and bone pain. In recent years there has been an increase in evidence-based research surrounding *jamu*, scientifically proving some of its potential as a true medicine (see Table 5) and lifting it to daily modern clinical practice^25^. With its popularity and the fact that it can be sold on the self, several *jamu* manufacturers have added corticosteroids in their product, improving its potency and potentially increasing the chance of steroid overdose (Cushing syndrome, and ironically bone loss and osteporosis)^26,27^. It should be noted that often, *jamu* are not covered by the BPJS, but it is still affordable compared to modern medicines. The utilization of *jamu* besides modern treatment in Indonesia may become an example of how traditional medicine, backed by enough evidence, can play an important role in modern medical treatment.

Another main issue surrounding treatments are compliance – which is a complex issue worthy of its own independent research. Majority of Indonesians pay attentions to symptoms, when the symptoms disappear, Indonesian will be less motivated to continue their treatment. This is especially true for mobility health problems, where the main complaints are usually pain and discomfort when moving. Utilization of awareness campaign in multiple mass media might help increase awareness, but the experts also think that involvement of caregivers will show increase in compliance. Indonesian are very communal with strong ties towards their family and community^28^; involving the patient’s immediate surrounding towards their treatment will surely increase the patients compliance^29,30^.

The experts had also identified gaps in the provision of treatment and prevention of mobility health problems in Indonesia, which includes: unreliable screening methods, expensive diagnostic tests, lack of research, lack of public knowledge on certain mobility-related diseases, and the fact that the general public still concentrates on curative rather than preventive measures. Even though there are no silver bullets for all these issues, these realizations will help decision-makers tackle mobility health problems in Indonesia.

### Awareness Campaign

Even though the usage of apps for screening might be ineffective for the elderly, experts found that the elderly in Indonesia intensely utilizes social media and its messaging services. Facebook, Instagram, and WhatsApp groups play a pivotal role in the spread of health-related information. For better or worse, the role of social media had been especially prominent in the pandemic, e.g. personal information of COVID-19 patient zero in Indonesia became viral in social media without the patient’s consent in a matter of hours^31^. When used correctly, social media can become a robust awareness campaign tool as it is affordable, easily accessible and can transcend vast distances in just a matter of seconds^32^.

An interesting point taken from the expert interview is the utilization of religious activity and gathering as a hotspot for identifying and promoting awareness and treatment towards mobility health. With Muslims praying five times each day, there is a repetition of movement that bounds to highlight any mobility issue presents in an individual. Although unorthodox, this inadvertent screening method can be an easy and affordable way to identify mobility health issues in Muslims. Religious gatherings had also been identified as a spot for sharing information regarding mobility health; this includes spreading information regarding mobility problems and diseases and promoting treatments (more often herbal treatments) to alleviate said problems.

Health care professionals should pay more attention to these unconventional means of health promotions, as both the general population and health care professionals may gain mutual benefits from it. Appropriate, targeted, applicable, and effective health promotion programs and messages can be effectively conveyed in religious gatherings which offers large potential audiences^33^. In recent years, the Indonesian government had effectively applied a similar move with the involvement of religious groups to increase vaccinations uptake^34^.

### Supplements for Mobility Health

In choosing supplements, Indonesian tends to choose based on popularity instead of the supplement’s ingredients or proven benefits. Both standard and herbal supplements are popular, but they are more commonly used to support an ongoing treatment instead of preventing mobility health problems from occurring (see Figure 1). The use of fortified foods is common, especially those for improving bone health and preventing osteoporosis.

An analysis of supplement consumption behaviour of 21,923 adults found that supplementary consumption for disease prevention was high among healthy people living a healthy lifestyle. Healthy individuals might be the demographic that the expert misses as they are all healthcare professionals who would typically deal with an individual with a health problem. The study continues by recommending that local health care providers should actively develop a policy towards the uses of dietary supplements, which includes the risk of side effects and clearly describes the group of people who would most likely benefit from the supplement^35^. These changes would likely benefit the general public and increase appropriate supplements consumption to increase mobility health.

There seems to be a missed opportunity in utilising fortified food or milk (and nutrition in general) to increase mobility health. Consumption of fortified milk is common in Indonesia; further utilisation of such products for mobility health would be a practical solution. Usage of an advertisement on the product to properly educate the public on mobility health may bring benefits for both the producer and consumers. It is of no doubt that fortified food or milk had numerous benefits in preventing mobility health problems as well as improving the outcome of treatments^36–38^. With its familiarity and proven benefits, fortified food or milk should be used in the strategy of increasing awareness on mobility health.

### Limitation of the study

All experts for this study come from Java, an island filled with the country’s most advanced urban centres with easy access to medical care and health products. Their experience might not be relatable to others in less developed or rural areas of Indonesia, where mobility health is arguably more abundant due to many manual labour workers while health care resource is limited. It is also worth mentioning that while the paper aims to gather information from experts, an all-inclusive analysis that includes patients and the general population might help shed more light on mobility health in Indonesia. The researcher hopes that this input can be a backbone for improvement in future studies.

## CONCLUSION

Due Indonesian’s ageing population, attention has continuously increased on mobility health. Even though modalities for diagnosis are abundant, expensive prices for specific examinations hinders the general population from access to proper screening and diagnosis. Treatment of mobility health with herbal medicine (*jamu*) is increasing in popularity and might be beneficial if properly researched. Fortified food or milk can be utilized as means of increasing mobility health. Social media can be a promising tool to increase awareness and disseminate information regarding mobility health. In general, there needs to be a change in mindset from curative to prevention in both health care providers and the general population.

## Data Availability

All data produced in the present study are available upon reasonable request to the authors

## Author Contributions

R.W.B, and T.S conceptualized and designed this qualitative study, whereas M.Y, R.W.B, T.S and B.M.M contribute to the content of this study to enhance and make it clearly described in the manuscript under current and scientific context.

## Institutional Review Board Statement

Ethical clearance was obtained from medical faculty of Atmajaya University, Indonesia (NO: 13/11/KEP-FKIKUAJ/2020).

